# Medication-Wide Association Study of Alzheimer’s Disease and Related Dementias: Identifying Drug Candidates from Electronic Health Records through Explainable AI

**DOI:** 10.64898/2026.06.02.26354752

**Authors:** Yijun Shao, Ying Yin, Yan Cheng, John E. McGeary, Tracey H. Taveira, Debby W. Tsuang, Mark W. Logue, Siamack Ayandeh, Ali Ahmed, Edward Zamrini, Qing Zeng-Treitler

## Abstract

**Objective:** Alzheimer’s disease (AD) is a leading cause of death and disability, and treatment options for Alzheimer’s disease and related dementias (ADRD) remain limited. We applied a data-driven, mechanism-agnostic Medication-Wide Association Study Plus (MWAS+) framework to identify candidate medications associated with ADRD using longitudinal electronic health record data and explainable artificial intelligence (AI).

**Methods:** We used Veterans Health Administration electronic health record data from January 1999 to May 2022. The initial study population comprised 8,424,715 Veterans aged 65 years or older. Cases were defined by ADRD-related diagnosis codes or ADRD-related medication prescriptions, and controls were free of ADRD diagnosis and ADRD-related medication use. After exclusions and matching on sex, race, age at first encounter, and duration of follow-up, the primary analytic cohort included 505,817 matched case-control pairs (1:1; 1,011,634 Veterans). Longitudinal features were extracted from historical data up to 1 year before the index date and aggregated into 1-year intervals. We developed an upgraded Hybrid Value-Aware Transformer (HVAT 2.0) to jointly learn from longitudinal and nonlongitudinal clinical data while incorporating numerical values associated with clinical concepts, including cumulative medication dose. To enhance interpretability, we applied a medication-specific impact score method to estimate model-derived associations between medication exposure and ADRD risk.

**Findings:** The model demonstrated stable performance across data partitions, with area under the receiver operating characteristic curve values of 0.791 in the training set, 0.772 in the validation set, and 0.775 in the testing set. Metolazone and varenicline were identified as the top 2 candidate medications with negative impact scores, suggesting potentially protective associations with new-onset ADRD. The impact score was -0.196 per unit of cumulative dose for metolazone (1800 mg) and -0.134 per unit for varenicline (280 mg). Although individual-level impact scores varied, most exposed patients had negative scores, including 12,020 of 12,480 metolazone users (96%) and 8,341 of 8,786 varenicline users (95%).

**Implications:** This study demonstrates the feasibility of combining a medication-wide association framework, longitudinal dose-aware modeling, and explainable AI to identify candidate medications for ADRD from real-world electronic health record data. The findings should be interpreted as signals for hypothesis generation rather than evidence of causality. This framework may support prioritization of repurposing candidates for expert review, follow-up cohort validation, and future clinical investigation.

## 1. Introduction

Alzheimer’s disease (AD) is a leading cause of death and disability, affecting millions of individuals in the United States and worldwide.^1^ As the population ages, the number of adults aged 65 years or older living with AD is projected to increase from 6.7 million in 2023 to 13.8 million by 2060. The economic burden is substantial, with estimated annual costs of $345 billion for health care, long-term care, and hospice services in 2023, plus an additional $339.5 billion in unpaid caregiving.^1^ Despite this growing burden, treatment options remain limited.

Clinically, AD is often indistinguishable from AD and Related Dementias (ADRD),^2^ a broader category which includes related dementias such as vascular dementia, which share overlapping risk factors and overlapping symptoms, especially in the later stages. Biomarkers for in vivo biological confirmation were only recently starting to be obtained and developed; even then, not adequately, and their performance is not yet optimized. Management of ADRD includes both risk reduction and treatment. Risk reduction focuses on adopting health promotion and morbidity prevention methods, as outlined in the Lancet Commission,^3^ whereas treatment consists primarily of symptomatic therapy and, for biologically confirmed AD, disease-modifying therapy. No available therapy can reverse or definitively halt disease progression. Although novel pharmacologic therapies have emerged for the treatment of ADRD, their clinical benefits remain modest.^4-6^

Traditional drug discovery and clinical development require years of preclinical and clinical testing. These challenges have increased interest in repurposing existing drugs for ADRD. Several agents, including statins,^7^ antihypertensives,^8^ nonsteroidal anti-inflammatory drugs (NSAIDs),^9^ pioglitazone,^10^ and glucagon-like peptide-1(GLP-1) agonists,^11^ have been investigated in ADRD, with observational studies suggesting possible associations with reduced dementia risk. Many prior repurposing efforts have leveraged existing biological knowledge, including targeting amyloid-β aggregation, tau phosphorylation, neuroinflammation, metabolic dysfunction, and vascular pathways. However, randomized trials for repurposed drugs have largely yielded negative or inconclusive results, potentially to the heterogeneity of ADRD pathology.^12-16^ To date, no repurposed agent has demonstrated definitive disease risk-modifying efficacy in ADRD.

In contrast, this study, rather than use prior biological knowledge, we used a data-driven, mechanism-agnostic approach within a broader initiative utilizing a paradigm called the Medication-Wide Association Study Plus (MWAS+) framework.^17^ Hypotheses generated during the MWAS phase are subsequently tested and validated in the “plus” phase. This report focuses on the first stage of MWAS+, hypothesis generation; candidate drugs identified through this process are subsequently evaluated through systematic expert review and follow-up cohort studies.

## 2. Methods

This study included cohort assembly, feature extraction, AI model development, and model explanation for candidate drug identification. We used a large national electronic health record (EHR) dataset from the Veterans Health Administration (VHA), comprising more than 20 years of longitudinal clinical data. This resource enables evaluation not only of drug initiation, but also of cumulative exposure and its relationship to clinical outcomes.

The AI model used an upgraded design of the original Transformer-based architecture called the Hybrid Value-Aware Transformer (HVAT).^18^ The Transformer architecture has become a leading deep neural network (DNN) architecture for learning from sequence data, revolutionizing the field of natural language processing and being the foundational technology for most of today’s large language models such as, including GPT, Gemini, and Llama. This success has motivated researchers to explore its application in healthcare. Despite the similarities between longitudinal clinical data and natural language data, clinical data presents unique complexities that make adapting the Transformer architecture to this domain challenging, including heterogeneous modalities, irregular temporal structure, and associated numerical values. To address the complexities, we designed HVAT, which can jointly learn from longitudinal and non-longitudinal clinical data and specifically leverage numerical values (e.g., lab values) associated with clinical codes/concepts (e.g., lab tests) from the longitudinal data. For this study, we had to upgrade the original HVAT design to correctly utilize the cumulative dose values associated with medications. The upgraded version is referred to as HVAT 2.0.

The machine learning models capture nonlinear relationships and accommodate a high-dimensional covariate space. However, such models, particularly DNNs, are often criticized as “black boxes” due to their complexity and large number of parameters. To enhance interpretability, we applied the impact score assessment, a novel explainable AI (XAI) method developed by our team, to support robust identification of potential drug candidates.^19^ We focused on medication-specific impact scores for ADRD risk (i.e., those medications with the strongest potentially protective or risk-conferring associations). Because the model incorporated cumulative medication dose, the XAI method also enabled estimation of dose-response patterns for included medications.

### 2.1 Data Source, Study Design, and Cohort Assembly

The primary data source for this analysis is the VHA Corporate Data Warehouse (CDW). A case-control study design was used, with patients with ADRD as cases and those without ADRD as controls. The initial study population comprised 8,424,715 Veterans aged 65 years or older from January 1999 to May 2022. Cases were defined as patients identified as having ADRD based on either ADRD-related diagnosis codes **(eTable 1)** or ADRD-related medication prescriptions **(eTable 2**). Patients with early-onset dementia, defined as those younger than 65 years at the time of first ADRD diagnosis, were excluded because of potential differences in disease etiology and clinical presentation. To improve case ascertainment, patients were also required to have at least 2 distinct ADRD-related encounters.

Controls were defined as patients with no ADRD diagnosis and no ADRD-related medication prescriptions by the time of their last VA visit or as of May 2022. For both cases and controls, patients with fewer than 5 years of follow-up from the first encounter date to the last follow-up date were excluded because of insufficient longitudinal medical history. For cases, the last follow-up date was defined as the first ADRD encounter date; for controls, it was defined as the last encounter date. Patients with diagnoses resembling, but distinct from, ADRD, such as senile delusion, were also excluded. The ICD codes used for these exclusions are listed in **eTable 1**. After these exclusions, 5,505,256 eligible Veterans remained, including 422,962 first identified through ADRD diagnosis and 83,293 first identified through ADRD medication use (**Figure 1**).

**Figure 1:**
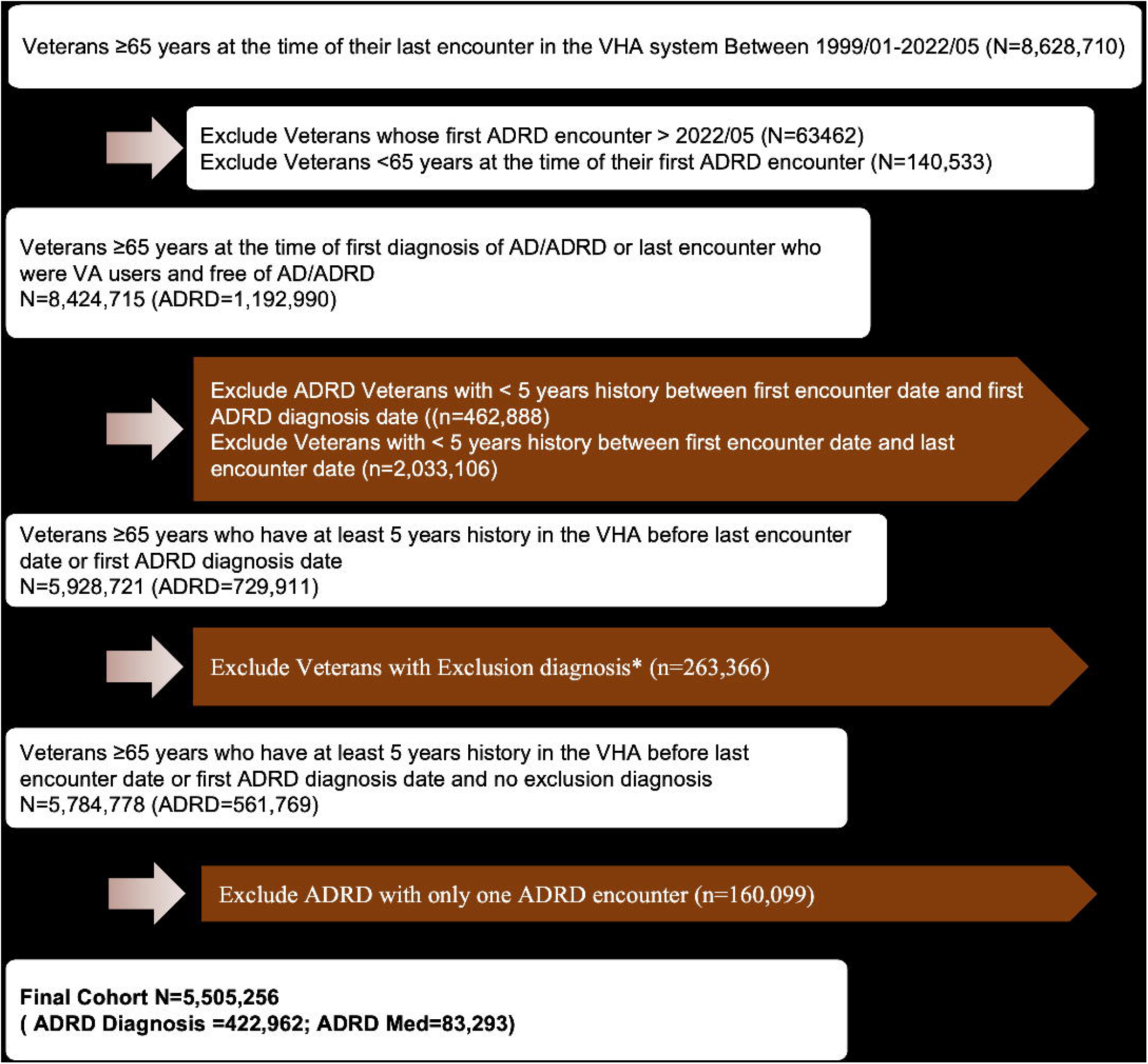
Study cohort.

Controls were then matched to cases on sex, race, age at first encounter in CDW, and the first encounter within 6 months to ensure comparable demographic characteristics and timing of entry into the health system. The index date for each matched pair was defined as the date of the case’s first ADRD diagnosis. Controls were required to have at least 12 months of ADRD-free follow-up after the index date within the electronic health record system to confirm the absence of ADRD at the index date. A total of 505,817 matched case-control pairs were identified for analysis, representing 1,011,634 Veterans in the primary analytic cohort.

### 2.2 Feature Extraction

For each individual, features were extracted from all available historical data up to 1 year before the index date. Data from the year immediately preceding the index date were excluded to avoid capturing prodromal or early manifestations of ADRD, including changes in symptoms, clinical evaluation, or treatment patterns that may precede formal diagnosis. Including this period could bias the model toward short-term predictors of near-term diagnosis rather than longer-term associations with medication exposure. The observation period was divided into 1-year intervals, and features were aggregated within each interval to construct a temporally ordered sequence. Natural numbers 1, 2, 3, … were used to order the 1-year intervals backwards from the latest to the earlier. These numbers were referred to as temporal indexes.

#### 2.2.1 Medication Feature Construction

Medication histories were obtained from the outpatient pharmacy domain of the Veterans Affairs (VA) Corporate Data Warehouse. Because the analysis focused on long-term medication exposure, we included oral medications with a prevalence greater than 1% in the analytic cohort; insulin was also retained because of its common use as a chronic maintenance therapy. Insulin products were categorized as rapid-acting, short-acting, intermediate-acting, long-acting, or combination formulations. Compound medications were separated into individual active ingredients using regular expression parsing, yielding 243 unique medications.

Dose strength was normalized to a common unit for each medication. Prescriptions were assigned to yearly exposure intervals based on dispensing date rather than actual days of use. Consequently, prescriptions filled near the end of a given year could be allocated to that year even if some of the supplied medication was used in the following year.

Annual cumulative exposure was calculated as dispensed days’ supply multiplied by normalized dose strength. To reduce the influence of implausible values likely attributable to data entry or dispensing errors, daily dose and quantity dispensed were capped at 3 times the 95th percentile for each medication, stratified by dose strength. Medication exposure within each interval was quantified as cumulative dose and summed across prescriptions and refills.

#### 2.2.2 Other Longitudinal Features

Other clinical data, including diagnoses, procedures, and note titles, were aggregated into 1-year intervals. For diagnosis features, ICD-9-CM codes were first mapped to ICD-10-CM codes using the Centers for Medicare & Medicaid Services General Equivalence Mappings, after which codes were grouped into 203 ICD-10-CM code blocks. For each occurrence of the code block, we assigned a value of 1, and aggregated the values using sum, so that for each time interval, the code blocks had counts as values. Features with a prevalence greater than 1% in the analytic cohort were retained. Selected laboratory tests were also included, and values were aggregated within each interval using the median. All model features are listed in **eTable 3**.

### 2.3 AI Modeling

#### 2.3.1 HVAT 2.0

The HVAT 2.0 architecture was an upgrade from the original design and the method of preprocessing the longitudinal data for the HVAT model was also modified from the original design accordingly (**Figure 2**).^18^ This upgrade was necessary because we realized there were two types of numerical values in the longitudinal data, while the original HVAT was only suitable to handle the first type.

**Figure 2.**
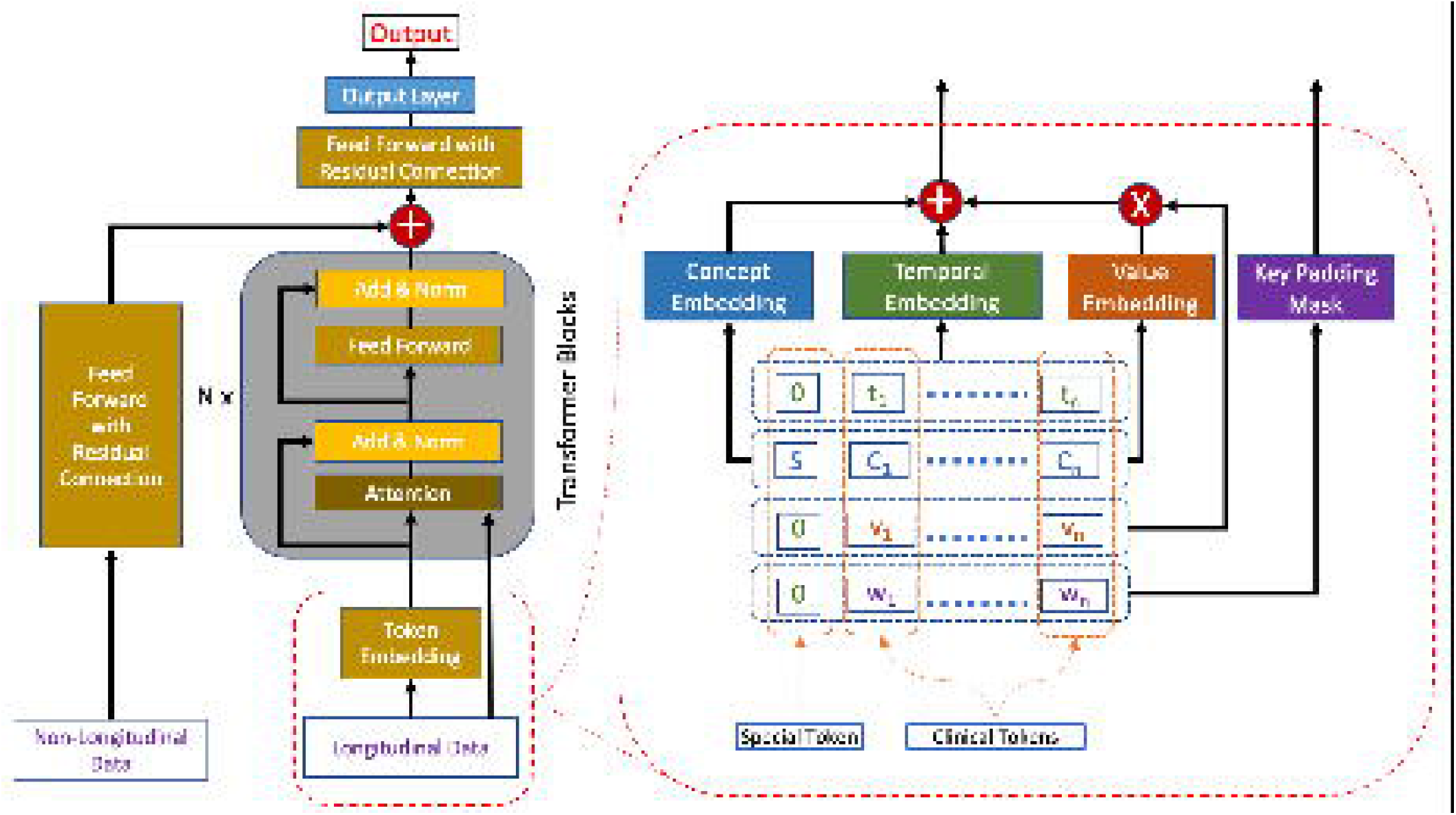
Hybrid Value-Aware Transformer (HVAT) 2.0 architecture. The left panel shows the full architecture. The box outlined with red dashed lines on the right provides an enlarged view of the smaller red dashed box on the left. The 2 arrows extending from the enlarged box correspond to the 2 arrows connecting the smaller box to the Transformer blocks

##### 2.3.1.1 Longitudinal Data Representation

The longitudinal data can be abstracted as a set of minimal yet semantically complete data elements, each containing at least three fields: a patient ID, a timestamp, and a clinical concept. The clinical concept may be from a vocabulary of standard terminology (e.g., ICD codes) or of a non-standard terminology (e.g., fitness). Some data elements may also have a fourth piece: a numerical value related to the concept. Whether the data element has a value usually depends on the type of concept it represents. For example, for concepts that are diagnostic codes (e.g., ICD codes), there are usually no values in the data elements; for lab tests or medications, the data elements usually include lab values from the lab tests or dose values from prescriptions or refills.

##### 2.3.1.2 Numerical Value Types and Aggregation

A key feature of our framework is the distinction between two types of numerical values in longitudinal clinical data (**Table 1**). The first type represents a patient’s internal physiologic state and includes observed or measured values, such as laboratory results and vital signs. The second type represents the magnitude of an external action, or intervention or health utilization, such as cumulative medication dose, visit count, or length of stay. Although these value types may appear similar in the raw record structure, they differ in meaning and therefore require different handling methods during preprocessing and model implementation.

**Table 1.** Comparison of the 2 Types of Numerical Values in Longitudinal Clinical Data.

|  |  |  |
| --- | --- | --- |
| **Characteristic** | **Type 1: Internal Physiologic State** | **Type 2: External Action or Intervention** |
| Conceptual meaning | Represents a patient's internal physiologic status | Represents the magnitude or intensity of an external action, intervention, or utilization |
| Typical source | Observation or measurement | Treatment, therapy, or health care utilization |
| Examples | Hemoglobin A1c, body temperature, laboratory values, vital signs | Cumulative medication dose, outpatient visit count, inpatient length of stay |
| Clinical interpretation | Reflects a measured patient state at a given time | Reflects accumulated exposure or intensity of an action |
| Appropriate aggregation within a time interval | Mean, median, minimum, or maximum | Sum |
| Why sum is or is not appropriate | Sum is not meaningful because the value should remain a plausible observed measurement | Sum is appropriate because the value represents cumulative exposure or utilization |
| Interpretation of absence of value | Absence of a value does not necessarily imply value was zero; zero may itself be a meaningful value | Absence of a value is typically equivalent to a value of zero, meaning no action or intervention or utilization occurred |
| Model handling in HVAT | Entered as normalized continuous values through value embedding | Transformed and incorporated through the key padding mask and processed in the attention modules |
| Normalization approach | Standardized within concept, typically to mean 0 and standard deviation 1 | Scaled by a high percentile value, then log-transformed |

At the data preprocessing stage, each patient’s longitudinal history was divided into equal-length time intervals. Within a given interval, multiple data elements could share the same concept but differ in timestamp; these elements were aggregated into a single feature for feature extraction. When repeated elements for the same concept also contained numerical values, those values were further aggregated into a single summary value. The aggregation method differed according to the type of numerical value, as shown in **Table 1**.

##### 2.3.1.3 Encoding of Numerical Values

The method for processing values of the first type is given in the original paper.^17^ In contrast, the values of the second type required a different approach at the model processing stage. A key property of these values is that, when present, they are always strictly positive (i.e., > 0), and the value range can be extended to include 0, which is commonly understood as “no intervention”. whereas in reality, “absence of the corresponding intervention” is mostly represented by the absence of a record of the “intervention” in the record rather than by a recorded value of 0 for the intervention. For example, nonuse of a medication is typically reflected by the absence of a medication in the record, not by a dose value of 0. In contrast, values of the first type do not have this property, because 0 may represent a valid observed measurement. This distinction required the two value types to be handled differently in the HVAT model.

The HVAT architecture includes an attention mechanism that processes sequences of clinical tokens with optional masks. There are two types of masks: the key padding mask (KPM) and the attention mask, but we will only use the KPM. The original purpose of the KPM is to inform the model which tokens in a sequence are not real but padding tokens to make the sequence align with the other sequences in length in a batch of sequences for batched learning. Usually, the KPM takes binary values: 0 and –∞, with 0 indicating a real token while –∞ indicating a padding token. The HVAT model will ignore the tokens with KPM = –∞. We realized that thec KPM values can be extended to any decimal values, while the meanings of the two values 0 and –∞ are preserved. Note that the HVAT model output varies continuously along with the KPM values, and as a KPM value tends to –∞, the model output tends continuously to the one corresponding to the KPM value being –∞, or corresponding to the token being deleted from the sequence. This is crucial for the XAI method to work for this study.

For values of the second type, we assumed that all observed values were positive and that smaller values had effects closer to the absence of values. For each concept with the values of the second type, we first normalize the values through division by a value that is sufficiently large but not exceeding the maximum (e.g., the 95^th^ percentile), so that most of the normalized values lie between 0 and 1. This way of normalization is more robust than simply dividing by the maximum because the maximum could be an outlier (much larger than most of the other values). Next, we take the natural logarithm of the normalized values so that most of them become negative values. Then these values are used as the KPM values to be received by the HVAT model. Since log(0) = –∞, for raw values very close to 0, the KPM values are very close to –∞, and the effects are close to the token being ignored by the model, which fulfills our purpose.

##### 2.3.1.4 Clinical Token Construction

The specific method for data pre-processing (feature extraction) is described in the previous section, followed by the value processing steps described above.

A clinical token is a quadruple *(t, C, v, w)*, where **t** is the temporal index, *C* is the concept, *v* is a value of the first type, and *w* is a value of the second type. From data elements without values, we generated tokens with *v* = 0 and *w* = 0. From data elements with values of the first type, we generated tokens with v equal to the normalized original value and *w* = 0. From data elements with values of the second type, we generated tokens with *v* = 0 and *w* equal to the values processed as described above.

The method for normalizing values of the first type is not unique. In this study, we used standard normalization by subtracting the mean and dividing by the standard error, such that the normalized values had mean 0 and standard error 1. This was done separately for each concept *C*. Thus, for each patient, we generated a set of clinical tokens from the longitudinal data. We added a special token, *(0, S, 0, 0)*, to every patient for summarization purposes, where *S* is an artificial concept defined to be different from all other concepts.

Specifically in this study, the concepts for which data elements had no values were procedures (represented by CPT codes) and note titles; the concepts for which data elements had values of the first type were the selected laboratory tests; and the concepts for which data elements had values of the second type were medications and ICD-10-CM code blocks. The values for medications were cumulative doses, and the values for the code blocks were counts. For cumulative dose values, normalization used the 95th percentile, whereas for code block counts, normalization used the 99th percentile. For the temporal indices, we ordered the time intervals backward (i.e, from latest to earliest) and assigned temporal indices 1, 2, 3, and so forth in that order.

For procedures and note titles, we selected 400 unique procedures and 400 unique note titles with the highest discriminatory power for outcome prediction. We used the selection method described in the original paper.^17^ For laboratory tests, we selected 10 features: calcium, cholesterol, creatinine, hemoglobin A1c (HbA1c), high-density lipoprotein (HDL), low-density lipoprotein (LDL), potassium, sodium, triglycerides, and vitamin D. For medications, we used 243 unique medications, and for diagnosis features, we used a total of 203 unique code blocks. In total, the model input included 1256 unique concepts.

The nonlongitudinal data included age, sex, race, and ethnicity. Age was a continuous variable, and its values were normalized to have mean 0 and standard error 1. Sex was a binary variable. Ethnicity was a categorical variable with 3 categories: (1) Hispanic, (2) non-Hispanic, and (3) unknown. Using non-Hispanic as the reference category, we converted ethnicity into 2 binary variables: Ethn_Hispanic and Ethn_Unknown. Race was a categorical variable with 7 categories: (1) White, (2) Black or African American, (3) Asian, (4) American Indian or Alaska Native, (5) Native Hawaiian or Pacific Islander, (6) other race, and (7) unknown. Using White as the reference category, we converted race into 6 binary variables in a similar way. Overall, there were 10 nonlongitudinal variables.

##### 2.3.1.5 HVAT Architecture and Model Training

The HVAT model consisted of 2 Transformer blocks with an embedding dimension of 48. For each patient, the model generated a single output value, p, ranging from 0 to 1 and representing the predicted probability of case status. The model took as input a sequence of clinical tokens containing values v and w. The values v were incorporated through the value embedding defined in the original HVAT framework, whereas the values w were incorporated as key padding mask (KPM) inputs (**Figure 2**).

The cohort was divided into 3 subsets: training (80%), validation (10%), and testing (10%). Model performance was evaluated using the area under the receiver operating characteristic curve (AUC). Training was performed using minibatches of 50 patients and the Adam optimizer with a learning rate of 0.0001. After each epoch, AUC was evaluated on the validation set and model weights were saved. Training was stopped when validation AUC failed to improve for 10 consecutive epochs. The model weights corresponding to the best validation AUC were then reloaded to define the final trained model, which was subsequently evaluated on the testing set to obtain the final AUC.

#### 2.3.2 AI Explanation

To interpret the HVAT model, we calculated medication-specific impact scores, first at the individual level (i.e., for each Veteran), and then at the population level. For each medication and for each patient who had been on this medication, the individual-level impact score on the patient was defined as follows:

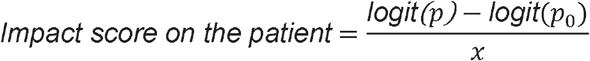

where logit 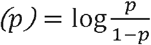 was the log odds, *X* was the sum of all the normalized cumulative dose values on the patient for this medication over all the temporal indices, *p* was the model output value with the patient’s data as the input, and *p*_0_ was the model output with all the tokens having this medication as the concept deleted from the patient’s longitudinal data (i.e., *p*_0_ was the model output with the same input data except the total cumulative dose of this medication for this patient was decreased to zero). Note that the individual-level impact score is not defined on the patients who had never been on the medication. The population-level impact score was just the mean of the individual-level impact scores for each medication.

We normalized the values in the denominator to make the impact scores of different medications comparable. Recall that the cumulative dose values were normalized using the 95^th^ percentile of the cumulative doses over all the year-long intervals across the entire cohort was used for normalization. This normalization also made the impact scores of different medications comparable to each other. Under this normalization, one unit is exactly the 95^th^ percentile. The impact score can be interpreted as the average change in log-odds of ADRD for each one unit increase in the normalized total cumulative dose of the medication.

Then we ranked the medications based on the (population-level) impact scores. Medications with the most negative impact scores were prioritized as candidates for further evaluation for ADRD prevention.

## 3. Results

### 3.1 Cohort Assembly

The analytic cohort consisted of 1,011,634 Veterans, including 505,817 case-control pairs matched for age, sex, race, and time of first encounter in the healthcare system (**Table 2**). The mean (SD) age at index was 80.3 (7.5) years. The cohort was predominantly male (990,952 [98.0%]) and White (763,952 [75.5%]), with 108,880 participants (10.8%) Black or African American. Ethnicity was Hispanic or Latino in 37,668 participants (3.7%) and not Hispanic or Latino in 881,295 (87.1%); ethnicity was unknown for 92,671 participants (9.2%).

**Table 2.** Demographic Characteristics of Cases and Controls at Index Date.

|  |  |  |
| --- | --- | --- |
| **Characteristic** | **Case, No. (%) or Mean (SD)** | **Control, No. (%) or Mean (SD)** |
| Total patients | 505,817 | 505,817 |
| Age at index, mean (SD) | 80.2 (7.5) | 80.3 (7.4) |
| **Sex** |  |  |
| Male | 495,476 (98.0) | 495,476 (98.0) |
| Female | 10,341 (2.0) | 10,341 (2.0) |
| **Race** |  |  |
| White | 381,976 (75.5) | 381,976 (75.5) |
| Black or African American | 54,440 (10.8) | 54,440 (10.8) |
| Other race or unknown | 69,401 (13.7) | 69,401 (13.7) |
| **Ethnicity** |  |  |
| Hispanic or Latino | 12,168 (2.4) | 25,500 (5.0) |
| Not Hispanic or Latino | 441,289 (87.2) | 440,006 (87.0) |
| Unknown | 52,360 (10.4) | 40,311 (8.0) |

### 3.2 Model Performance and Candidate Medication Identification

The final analytic sample included 1.01 million individuals. Model performance was stable across data partitions, with an area under the receiver operating characteristic curve of 0.791 in the training set, 0.772 in the validation set, and 0.775 in the testing set.

Model explanation identified metolazone and varenicline as the top 2 candidate medications with negative impact scores, suggesting potentially protective associations with ADRD outcomes. The impact score was −0.196 per unit (1800 mg) of cumulative dose for metolazone and −0.134 per unit (280 mg) for varenicline.

### 3.3 Patient-level Impact Analysis for Metolazone and Varenicline

Individual-level impact scores for both metolazone and varenicline were distributed across 0, but most were negative (**Figure 3**). On the log-scaled histograms, both drugs showed a left-skewed distribution, indicating that a small subset of patients had more strongly negative impact scores. Overall, these findings suggest that in most exposed patients, the model attributed metolazone and varenicline to lower predicted ADRD risk, although the magnitude of this association varied across individuals.

**Figure 3.**
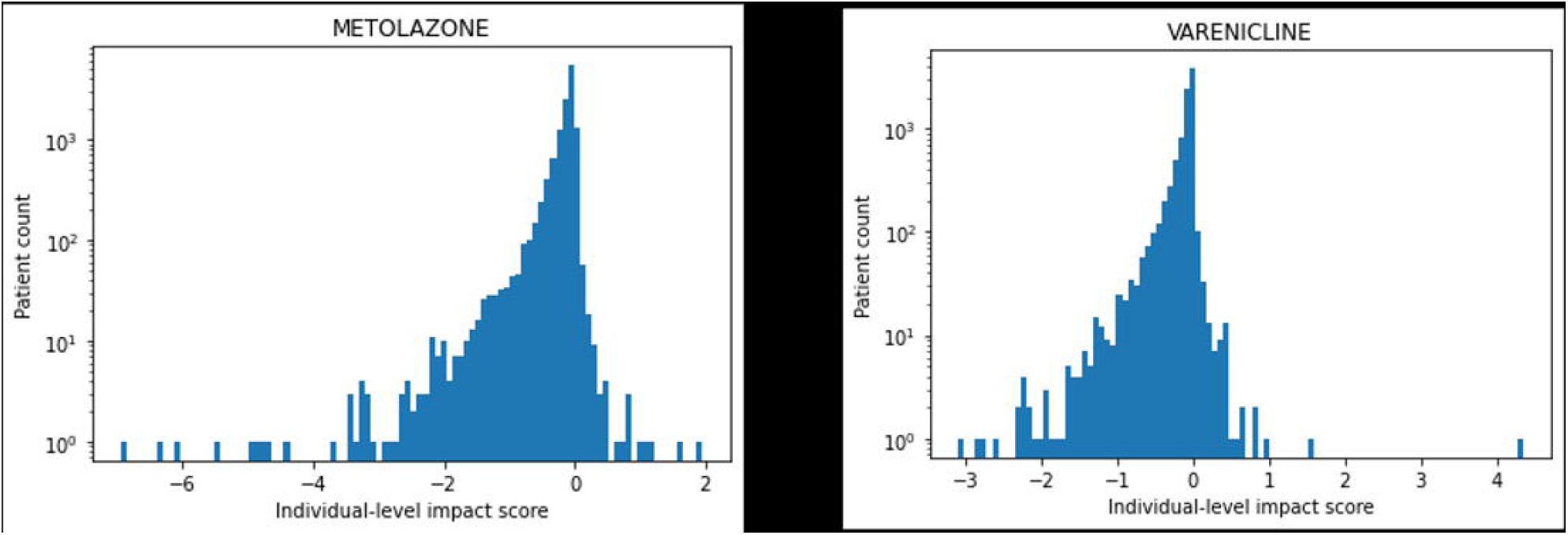
Histograms of individual-level impact scores with a log-scaled y-axis. A) impact scores of metolazone, B) impact scores of varenicline.

Among 12,480 metolazone users, 12,020 (96%) had negative metolazone impact scores and 460 (4%) had positive impact scores (**Table 3**) Compared with metolazone users with positive impact scores, those with negative impact scores were younger at the index date (mean age, 79.4 vs 81.5 years), and had a lower proportion of ADRD cases (44% vs 55%). Sex distributions were similar between groups. By race, the negative-impact group included a higher proportion of White patients (73% vs 65%) and a lower proportion of Black patients (16% vs 22%). Ethnicity distributions were similar between groups.

**Table 3.** Characteristics of Metolazone and Varenicline Users Stratified by Drug-Specific Individual-Level Impact on ADRD.

|  | **Metolazone user (N = 12,480)** |  | **Varenicline user (N= 8786)** |  |
| --- | --- | --- | --- | --- |
| **Characteristic** | **Negative metolazone impact on ADRD** | **Positive metolazone impact on ADRD** | **Negative varenicline impact on ADRD** | **Positive varenicline impact on ADRD** |
| Patient count | 12,020 | 460 | 8341 | 445 |
| Medication use duration, y, mean | 2.4 | 2.0 | 1.5 | 1.4 |
| ADRD, % | 44 | 55 | 47 | 26 |
| **Age at index date** |  |  |  |  |
| Mean | 79.4 | 81.5 | 72.9 | 76.7 |
| Standard deviation | 7 | 7.3 | 5.4 | 5.9 |
| **Sex, female, %** | 1 | 2 | 3 | 2 |
| **Race, %** |  |  |  |  |
| Black | 16 | 22 | 9 | 7 |
| White | 73 | 65 | 84 | 83 |
| Other | 11 | 14 | 7 | 10 |
| **Ethnicity, %** |  |  |  |  |
| Hispanic | 3 | 3 | 2 | 2 |
| Not Hispanic | 89 | 90 | 95 | 93 |
| Unknown | 8 | 7 | 4 | 5 |

Similarly, among 8786 varenicline users, 8341 (95%) had negative varenicline impact scores and 445 (5%) had positive impact scores, indicating that varenicline exposure was associated with a lower model-predicted ADRD risk in most exposed patients (**Table 3**). Compared with varenicline users with positive impact scores, those with negative impact scores were younger at the index date (mean age, 72.9 vs 76.7 years) and had a higher proportion of ADRD cases (47% vs 26%). Sex and ethnicity distributions were similar between groups. Racial differences were modest, with the negative-impact group including a slightly higher proportion of White patients (84% vs 83%) and Black patients (9% vs 7%) and a lower proportion of patients in the other-race category (7% vs 10%). The negative-impact group also had a slightly lower proportion of Hispanic patients.

## 4. Discussion

### 4.1 The MWAS+ Framework as a Hypothesis-Generating Approach

AD drug repurposing has a long history, with most prior efforts guided by specific mechanisms or preexisting hypotheses. For example, prior work has examined antihypertensives, anti-inflammatory agents, and metabolic drugs such as metformin^20^ on the basis of known or suspected biologic pathways. In contrast, our approach is hypothesis-free and mechanism-agnostic, leveraging large-scale observational data to identify candidate associations without prespecifying mechanisms. This strategy may reveal unexpected signals that would not emerge from traditional target-based approaches, although such associations require careful validation.

In this study, we assembled a large longitudinal cohort from the VA electronic health record system, enabling comprehensive analysis of cumulative medication exposure and ADRD outcomes. Using this dataset, we trained a deep learning model that demonstrated stable predictive performance. Our explainable artificial intelligence framework identified both potentially novel and previously suggested drug candidates. To our knowledge, this represents one of the first efforts to combine a medication-wide association framework, longitudinal EHR data, and explainable AI for hypothesis generation in ADRD drug repurposing.

### 4.2 Methodological Innovation of the HVAT Framework

An important innovation in these analyses was the use of value-aware methods. In clinical data, numerical values are important because some concepts have associated values, such as laboratory measurements and medication doses, whereas others do not. This differs from natural language data, in which information is primarily represented as unstructured tokens. Our approach explicitly incorporates both value-bearing and non–value-bearing concepts within a unified framework.

In addition, our framework distinguishes between different types of numerical values. Some values reflect a patient’s internal physiologic state, such as laboratory results, whereas others reflect the magnitude of an external action or intervention, such as cumulative medication dose. We believe this is an important methodological contribution because these value types differ in clinical meaning and should not be processed in the same way. This value-aware design allowed the model to preserve clinically meaningful information that might otherwise be lost.

### 4.3 Why We Studied ADRD Rather Than AD Alone

This study focused on ADRD rather than AD alone for both practical and clinical reasons. In routine EHR data, the diagnosis of AD is often difficult to distinguish reliably from other related dementias, especially in the absence of biomarker confirmation or autopsy. Accordingly, constructing an ADRD cohort allowed us to study a broader and more clinically realistic phenotype within the available data.

This broader approach may also capture medications that act through mechanisms relevant across multiple dementias rather than only one diagnosis within the ADRD spectrum. For example, some medications may exert more general benefits through anti-inflammatory, antioxidant, nootropic, cardiovascular, or cerebrovascular effects. Such mechanisms may be relevant across the broader ADRD population. At the same time, a medication that works primarily for AD could still emerge as a signal within an ADRD-based analysis if its effect on AD is sufficiently strong, even if that signal is diluted by inclusion of other dementia subtypes.

Thus, although our approach is broad, it can still generate clinically meaningful insights. Signals identified in ADRD can inform subsequent investigation of the mechanism, target population, and disease specificity. In this sense, the present strategy may serve as a first-pass screening approach, with future follow-up studies examining whether identified signals are more relevant to AD specifically or to ADRD more generally.

### 4.4 Interpretation of MWAS+ Findings as Signals Rather Than Causal Effects

The findings from this MWAS+ framework should be interpreted as signals or associations rather than as evidence of causality. This approach is intended for hypothesis generation, not for immediate clinical recommendation. The fact that a medication has a favorable impact score does not mean that it should be used broadly in patients with ADRD. Rather, these findings provide initial insights that require additional interpretation in the context of pharmacology, clinical experience, the existing literature, and further research.

Some identified associations may appear biologically plausible, whereas others may be less intuitive. In either case, the next step is not to assume benefit, but to evaluate the strength, specificity, and clinical plausibility of the signal. This process may support more targeted follow-up studies, including expert review, replication in independent datasets, refined observational analyses, and, where justified, clinical trials. In that sense, this work should be viewed as a starting point. The goal is not to provide definitive answers, but to identify candidate signals that may ultimately lead to repurposable therapies.

### 4.5 Candidate Medications Identified

Notably, metolazone emerged as a potentially novel candidate, while varenicline has been suggested in prior studies.^21^ The mechanistic pathways of metolazone and varenicline will be discussed in other manuscripts. A larger set of candidate medications has been identified and is currently undergoing systematic evaluation through expert panel review.^22^

### 4.6 Limitations

Several limitations should be considered. First, the VA population may limit generalizability due to its demographic composition. Second, ADRD phenotyping remains imperfect, as it relies primarily on diagnosis codes and medication proxies rather than biomarker confirmation. Third, ADRD is a heterogeneous condition, which may obscure subtype-specific associations. Finally, EHR-derived features are subject to data quality limitations, including inaccuracies in coding (e.g., ICD-based comorbidity definitions) and missing or inconsistent data.^23^ Additionally, this analysis was limited to oral medications and therefore did not include most GLP-1–based therapies. Moreover, medication exposure was inferred from dispensing records rather than confirmed medication use.

Nevertheless, these AI-informed MWAS analyses may prove to be a useful new lens to identify medications for ADRD that are urgently needed.

### 4.7 Future Directions

Our future work will focus on continued expert panel validation of identified candidates, prospective cohort validation, and replication in non-VA EHR datasets to assess generalizability and robustness. The concurrent and future analyses will further characterize the expert review framework, including the clinical, pharmacologic, and methodological considerations underlying candidate prioritization, and will also evaluate the translational potential of high-priority medications identified through these analyses.

## Funding source

Research reported in this publication was supported by the National Institute on Aging (R01AG073474) and the Department of Veterans Affairs (I01BX006962-01A1). The views and conclusions expressed in this manuscript are those of the authors and should not be construed as representing the official policies of the National Institutes of Health or the Department of Veterans Affairs, either expressed or implied.

## Disclosure of Interest

The authors declare no conflicts of interest.

## Data Availability

The data analyzed in this study were obtained from the VA CDW. These data are accessible through the Veterans Affairs Informatics and Computing Infrastructure (VINCI), a secure computing environment located behind the VA firewall. Access to CDW data is restricted to authorized users and requires appropriate approvals. All data use is governed by VA policies and regulations regarding data security, privacy, and responsible data stewardship.

## Statement of AI Assistance

This manuscript was reviewed for grammar, syntax, and language clarity using ChatGPT (OpenAI, GPT-5.4 Thinking). The tool was used solely to support language refinement. All scientific content, interpretation, and conclusions were developed and verified by the authors.

## Ethical Approval

The study was approved by the Institutional Review Board of the Washington DC Veterans Affairs Medical Center.

## Acknowledgement

This work was conducted in the “AWS VA Enterprise Cloud Prospect” enclave, and we would like to thank Dr. Siamack Ayandeh for the creation of the analytics study mart environment and the VHA Office of the Research and Development for funding of the Cloud Credits.

## Notes

### Competing Interest Statement

The authors have declared no competing interest.

