## Supplementary material for "Medication-Wide Association Study of Alzheimer’s Disease and Related Dementias: Identifying Drug Candidates from Electronic Health Records through Explainable AI": Online tables

| **eTable 1. ICD Diagnosis Codes Used for ADRD Cohort Assembly** | | |
| --- | --- | --- |
| **Condition** | **ICD-9 CM** | **ICD-10 CM** |
| ADRD ICD | 290.0x, 290.10x, 290.4x, 291.2x, 292.82x, 294.10x, 294.11x, 294.20x, 294.21x, 294.8x, 331.0x, 331.19x, 331.2x, 331.7x, 331.82x, 331.89x, 331.9x, 780.93x | F01.50x, F01.51x, F02.80x, F02.81x, F03.90x, F03.91x, F10.27x, F10.97x, F13.27x, F13.97x, F18.17x, F18.27x, F18.97x, F19.17x, F19.27x, F19.97x, G30.0x, G30.1x, G30.8x, G30.9x, G31.09x, G31.83x, R41.2x, R41.3x |
| Exclusion ICD | 046.1x, 046.11x, 046.19x, 046.3x, 046.79x, 046.9x, 291.1x, 331.11x, 333.0x, 333.4x, 295x, 296.0x, 296.4x, 296.5x, 296.6x, 296.7x, 296.8x, 290.11x, 290.12x, 290.13x, 290.20x, 290.21x, 290.3x | A81.00x, A81.01x, A81.09x, A81.2x, A81.89x, A81.9x, F10.96x, G23.1x, G31.01x, G90.3x, F20x, F31x |

**eTable 2: Medication Used for ADRD Cohort Assembly**

**Medication Name**

Donepezil (Aricept)

Rivastigmine (exelon)

galantamine (Razadyne)

memantine (namenda)

| **eTable 3. Features Included in the Model** | | |
| --- | --- | --- |
| **Category** | **CDW Domain** | **Features** |
| Demographics | Patients | Patient demographic characteristics, including age, sex, race, and ethnicity |
| Medications | Outpatient pharmacy | ACARBOSE, ACETAMINOPHEN, ACETAZOLAMIDE, ACYCLOVIR, ALENDRONATE, ALFUZOSIN, ALLOPURINOL, ALPRAZOLAM, ALUMINUM HYDROXIDE, AMIODARONE, AMITRIPTYLINE, AMLODIPINE, AMOXICILLIN, APIXABAN, ASCORBIC ACID, ASPIRIN, ATENOLOL, ATORVASTATIN, ATROPINE, AZITHROMYCIN, BACLOFEN, BENAZEPRIL, BENZONATATE, BICALUTAMIDE, BISACODYL, BISMUTH SUBSALICYLATE, BUMETANIDE, BUPROPION, BUSPIRONE, BUTALBITAL, CAFFEINE, CALCITRIOL, CALCIUM, CALCIUM CARBONATE, CAPTOPRIL, CARBAMAZEPINE, CARBIDOPA, CARVEDILOL, CEFUROXIME, CELECOXIB, CEPHALEXIN, CETIRIZINE, CHLORPHENIRAMINE, CHLORTHALIDONE, CHOLECALCIFEROL, CILOSTAZOL, CIMETIDINE, CIPROFLOXACIN, CITALOPRAM, CLARITHROMYCIN, CLAVULANATE, CLINDAMYCIN, CLONAZEPAM, CLONIDINE, CLOPIDOGREL, CODEINE, COLCHICINE, COLESTIPOL, CYANOCOBALAMIN, CYCLOBENZAPRINE, DEXAMETHASONE, DEXTROMETHORPHAN, DIAZEPAM, DICLOFENAC, DICLOXACILLIN, DICYCLOMINE, DIGOXIN, DILTIAZEM, DIPHENHYDRAMINE, DIPHENOXYLATE, DIPYRIDAMOLE, DIVALPROEX, DOCUSATE, DOXAZOSIN, DOXEPIN, DOXYCYCLINE, DULOXETINE, ENALAPRIL, ERGOCALCIFEROL, ERYTHROMYCIN, ESCITALOPRAM, ETODOLAC, EZETIMIBE, FAMOTIDINE, FELODIPINE, FENOFIBRATE, FERROUS GLUCONATE, FERROUS SULFATE, FEXOFENADINE, FINASTERIDE, FISH OIL, FLUCONAZOLE, FLUOXETINE, FLUVASTATIN, FOLIC ACID, FORMOTEROL, FOSINOPRIL, FUROSEMIDE, GABAPENTIN, GEMFIBROZIL, GLIPIZIDE, GLUCOSE, GLYBURIDE, GUAIFENESIN, HYDRALAZINE, HYDROCHLOROTHIAZIDE, HYDROCODONE, HYDROXYZINE, IBUPROFEN, INDOMETHACIN, INSULIN_MIX, INSULIN_INTER, INSULIN_SHORT, INSULIN_RAPID, INSULIN_LONG, IRBESARTAN, ISOSORBIDE DINITRATE, ISOSORBIDE MONONITRATE, LACTOBACILLUS ACIDOPHILUS, LACTULOSE, LANSOPRAZOLE, LEVETIRACETAM, LEVODOPA, LEVOFLOXACIN, LEVOTHYROXINE, LIDOCAINE, LISINOPRIL, LOPERAMIDE, LORATADINE, LORAZEPAM, LOSARTAN, LOVASTATIN, MAGNESIUM CITRATE, MAGNESIUM HYDROXIDE, MAGNESIUM OXIDE, MECLIZINE, MEGESTROL, MELATONIN, MELOXICAM, METFORMIN, METHADONE, METHOCARBAMOL, METHOTREXATE, METHYLPREDNISOLONE, METOCLOPRAMIDE, METOLAZONE, METOPROLOL, METRONIDAZOLE, S_MULTIVITAMINS W/MINERALS , S_MULTIVITAMINS W/MINERALS, THERAPEUTIC , S_MULTIVITAMINS/MINERALS, THERAPEUTIC , MINOCYCLINE, MIRTAZAPINE, MISOPROSTOL, MONTELUKAST, MORPHINE, MOXIFLOXACIN, S_MULTIVITAMIN/OPHTH ANTIOXIDANT/LUTEIN , S_MULTIVITS W/MINERALS /CAP (NO VIT K), S_MULTIVIT/OPHTH AREDS2/LUTEIN/ZEAXANTHIN , S_MULTIVITAMIN , S_MULTIVITAMIN/OPTH AREDS SINGLE STRENGTH , S_MULTIVITAMINS , NAPROXEN, NIACIN, NICOTINE, NICOTINE POLACRILEX, NIFEDIPINE, NITROFURANTOIN, NITROGLYCERIN, NORTRIPTYLINE, NYSTATIN, OMEPRAZOLE, ONDANSETRON, OXYBUTYNIN CHLORIDE, OXYCODONE, PANTOPRAZOLE, PAROXETINE, PENICILLIN, PENTOXIFYLLINE, PHENAZOPYRIDINE, PHENYTOIN, PIOGLITAZONE, PIROXICAM, POTASSIUM CHLORIDE, PRAMIPEXOLE, PRAVASTATIN, PRAZOSIN, PREDNISONE, PREGABALIN, PRIMIDONE, PROCHLORPERAZINE, PROMETHAZINE, PROPRANOLOL, PSEUDOEPHEDRINE, PYRIDOXINE, QUETIAPINE, RABEPRAZOLE, RISPERIDONE, ROPINIROLE, ROSIGLITAZONE, ROSUVASTATIN, SALSALATE, SENNOSIDES, SERTRALINE, SILDENAFIL, SIMETHICONE, SIMVASTATIN, SODIUM BIPHOSPHATE, SODIUM PHOSPHATE, SOTALOL, SPIRONOLACTONE, SUCRALFATE, SULFAMETHOXAZOLE, SULINDAC, TAMSULOSIN, TEMAZEPAM, TERAZOSIN, TERBINAFINE, TESTOSTERONE, TETRACYCLINE, THEOPHYLLINE, THIAMINE, THROAT LOZENGE, TIOTROPIUM, TOLTERODINE, TOPIRAMATE, TRAMADOL, TRAZODONE, TRIAMTERENE, TRIMETHOPRIM, TRIPROLIDINE, VALACYCLOVIR, VALSARTAN, VARDENAFIL, VARENICLINE, VENLAFAXINE, VERAPAMIL, VITAMIN D, VITAMIN E, WARFARIN, ZOLPIDEM |
| Diagnoses | Outpatient, inpatient | ICD-9-CM and ICD-10-CM diagnosis codes grouped into ICD clusters based on GEM mapping |
| Procedures | Outpatient, inpatient | ICD procedure codes and CPT codes |
| Note titles | Outpatient | TIU standard note titles |
| Laboratory tests | Vital signs, labchem | BMI, serum calcium, serum cholesterol, hemoglobin A1c, HDL cholesterol, LDL cholesterol, serum potassium, serum sodium, triglycerides, and vitamin D |
| **Abbreviations:** BMI, body mass index; CPT, Current Procedural Terminology; GEM, General Equivalence Mapping; HDL, high-density lipoprotein; ICD, International Classification of Diseases; LDL, low-density lipoprotein; TIU, Text Integration Utilities. | | |
